# Knowledge, attitudes, and beliefs about sexual violence, and the implementation of sexual violence prevention programming: Survey of faculty at seven universities in Vietnam

**DOI:** 10.1101/2025.04.28.25326201

**Authors:** Daniel J. Whitaker, Quach Thu Trang, Meghan Macaulay, Tran Hung Minh, Xiangming Fang, Kathryn Yount

## Abstract

**Background:** College campuses are a common setting for sexual violence prevention efforts. Those efforts are often focused solely on students, though implementation theory suggests that campus faculty and leaders must be engaged for successful implementation. This is especially true in low- and middle-income countries, where resources are scarce and norms may support sexual violence. Little research has examined knowledge attitudes and beliefs around sexual violence and implementation readiness. We present findings from a faculty survey at seven Universities across Vietnam which assessed knowledge, attitudes, and beliefs, along with the acceptability and feasibility of sexual violence prevention programming.

**Method:** 2343 faculty (68% response rate) at seven Universities in Vietnam were surveyed preceding the implementation of the GlobalConsent intervention. Faculty reported on demographics; measures of knowledge, attitudes and beliefs (KAB) about sexual violence; and perceptions of the acceptability and feasibility for sexual violence implementation programming at their University. Analyses present descriptive data for key KAB measures, variation in KAB measures by key demographics, and regression models predicting implementation readiness.

**Results:** Faculty did not believe sexual violence was rare or problematic at their University, and while they tended not to endorse rape myths about victims, they tended to endorse rape myths about perpetrators, and beliefs supporting the need for sexual consent were moderate. Faculty did report positive campus climate for victims of sexual violence and believed sexual violence programming was acceptable and feasible. Female and younger faculty generally had more progressive mean scores for KAB measures. Several KAB measures related to the perceived feasibility and acceptability of sexual violence prevention programming, but the single strongest predictor of perceived feasibility and acceptability were perceptions of leader support for a positive campus climate around sexual violence.

**Conclusion:** Faculty perceived sexual violence prevention programming as both feasible and acceptable, and this was strongly related to university leadership’s support for a positive campus climate around sexual violence. Cultivating visible and consistent leadership support appears to be crucial to fostering faculty buy-in and enhancing prevention efforts. Interventions should address faculty KAB and actively engage institutional leaders; this is key to creating a supportive climate for victims of sexual violence.

## Overview of Sexual Violence

Sexual violence includes a range of behaviors comprising unwanted sexual contact, from sexual harassment to forcible rape, and includes any sexual activity when consent is not obtained or freely given (1). Sexual violence victimization is a public health problem in both its scope and consequences. In the U.S., over half of women and a third of men report some form of unwanted physical sexual violence, and about a quarter of women have experienced an attempted or completed rape (1). While men can be victims of sexual violence, over 90% of sexual violence crime victims are women (Rennison BJS report), and most sexual violence and unwanted sexual contact is perpetrated by young men. The consequences of being victimized from sexual violence are pervasive and well documented with profound adverse effects on physical and psychological health (2–6).

## Sexual Violence on College Campuses

College campuses have received a great deal of attention as a common setting for sexual violence perpetration, given that much sexual violence happens in young adulthood, and the elevated prevalence of alcohol consumption (7, 8). About one in five college women in the U.S. experience sexual violence, and international data suggest almost 18% of women report experiencing some kind of sexual violence (9). Campus-based interventions have been developed and tested with some success (10), and institutions of higher education have implemented prevention efforts to address campus sexual assault (11, 12). Those interventions have focused primarily on changing student knowledge, attitudes, beliefs, and behaviors related to sexual violence, either by directly targeting would-be perpetrators or by targeting young men to become active bystanders in preventing sexual violence (13, 14). Such bystander-based approaches attempt to engage students in prevention efforts by supporting shifts in prosocial bystander norms as well as proactive or reactive bystander behaviors that prevent others from perpetrating sexual violence (10, 15, 16). A recent review found that, while more rigorous studies are needed, the existing literature on bystander interventions is promising in that targeted mediators of sexual violence are affected in a positive manner (10, 17). For example, Park and Kim’s (17) meta-analysis found that bystander programs affected outcomes including knowledge and awareness of sexual violence, bystander efficacy, intention to intervene, and bystander behavior.

The student-focused approach to sexual violence prevention has been criticized as overly simplistic and reductive (18, 19), in that it fails to consider the full context of the college campus, including the perspectives and involvement of University leaders, faculty, and staff. Though a student-focused approach to prevention has been most prevalent, comprehensive theoretical models of sexual violence emphasize the influence of factors at multiple levels on sexually violent behavior, including inequitable gender norms and pro-violence social norms both within institutions and in the broader community environment (15, 20–25). There are increasing calls for prevention efforts to target levels beyond the student in an effort to affect real change to campus climate (26–29).

A clear need exists to engage University leaders and faculty in campus-based sexual violence prevention efforts. Those individuals help to create the overall culture and climate of the institution and control the allocation of resources to evidence-based programs to prevent sexual violence (19, 24, 26, 27, 29, 30). Engaging faculty and administrators in sexual violence prevention may change faculty knowledge, attitudes, and beliefs around sexual violence, which can contribute to an overall change in campus climate that makes sexual violence less likely.

Moreover, with increased awareness of sexual violence definitions, causes, and effects, faculty and staff can be better first-line resources for students when approached about an incident of sexual violence. These university employees may be more apt to recognize signs of traumatic events like sexual violence in students and may be more likely to act in an empathetic and resourceful way if a student discloses a personal experience of sexual violence.

Moreover, from an implementation perspective, engaging University leaders and faculty is critical for implementing and sustaining sexual violence prevention efforts, regardless of the primary intended recipients of those efforts. More generally, theory and research on implementation drivers have identified individual and organizational knowledge, attitudes, and beliefs as key drivers of successful implementation (31–34). Theories such as the Consolidated Framework for Implementation Research, or CFIR (32), and the Exploration, Preparation, Implementation, and Sustainment (EPIS) framework (35) include staff attitudes and organizational culture and climate as key variables that affect the success of an implementation. Research has demonstrated the importance of leadership engagement when implementing a new practice (36). When leaders are engaged and supportive, new practices are likely to be implemented, adopted, and sustained (36–38). Moreover, the attitudes from organizational leaders and staff about a practice are important in the implementation of that practice (34, 39). For college students, faculty and administrators represent the leaders of the University. The attitudes of faculty and administrators are likely to be important contextual determinants of student engagement in programming; if faculty and leaders do not support a student-directed program, students may be less likely to engage in that program and less likely to take it seriously.

## Sexual Violence in Vietnam

Most of the work on sexual violence prevention on college campuses has been conducted in high-income Western societies (9), and much less is known about sexual violence in low- and middle-income countries (LMICs) where societal norms and prevention resources are quite different. In Vietnam in 2019, an estimated 9.1% of Vietnamese women 15–19 years old and 18.0% of women 20–24 years old reported experiencing sexual violence since age 15 (40).

Reporting of sexual violence has also increased drastically in Vietnam in the past decade (40). In Vietnam, norms of masculine dominance remain widespread (22, 41, 42) and relationships in Vietnam remain age and gender hierarchical, with men and senior men in particular being privileged (41, 43). These norms contribute to attitudes, beliefs, and behaviors that support men’s sexual violent behavior. In qualitative research with undergraduates in Vietnam, gendered norms of masculine privilege, feminine discretion, and victim-blaming tended to normalize men’s sexually coercive behavior and undermine the concept of sexual consent (44, 45).

Thematic analysis of in-depth interview and focus groups of Vietnamese college students have revealed several myths and misconceptions about sexual violence, including beliefs that: rape results mainly from men’s uncontrollable sexual desire; victims provoke rape by their reckless behavior such as drinking or dressing provocatively; real rape is characterized by physical force from the perpetrator and fierce resistance on the part of the victim; and that rape occurs only under a narrow set of circumstances (45). Although young men and women have endorsed most myths equally, the justification of rape through victim blaming was much less common in young women’s narratives than in those of young men (45).

Given the prevailing norms supporting masculine dominance and sexual violence in Vietnam, it is critical that prevention efforts consider the full range of influences on student knowledge, attitudes, and behaviors when considering prevention programming. Yount and colleagues (29) conducted key informant interviews with university faculty and staff across Vietnam and found that informants named a range of factors consistent with theoretical implementation models as potential influences of sexual violence prevention programming. Key informants identified influences at the individual level (e.g., age, area of study facilitator knowledge), inner setting or University level (institutional culture, leader buy-in, acknowledgement of the problem of sexual violence), and outer setting or broader contextual factors (e.g., media, gender norms, policy). Thus, it is clear that prevention efforts must be considered in the broader context. For University-based prevention efforts, it is critical to understand faculty and leader attitudes about sexual violence, and the extent to which they see sexual violence prevention programming as acceptable and feasible.

## Overview of the Current Study

This paper reports on data from faculty employed at seven Universities in Vietnam who are taking part in *SCALE, Strategies for Implementing GlobalConsent to Prevent Sexual Violence in University Men*. SCALE is a national implementation trial of the GlobalConsent program (23). GlobalConsent is a web-based educational entertainment program designed to improve knowledge about sexual violence and consent, to reduce endorsement of harmful gender norms and rape myths, to increase empathy for the victims of sexual violence, to strengthen capacities for prosocial bystander intervention, and to promote bystander action as mechanisms to reduce sexually violent behavior among heterosexual or bisexual undergraduate men (15).

GlobalConsent was adapted from RealConsent (46), an on-line intervention for sexual assault prevention and alcohol education aimed at college-aged students. Evidence from the efficacy trial of GlobalConsent showed that it reduced sexually violent behavior and increased bystander action relative to a customized adolescent health attention-control condition (15). Moreover, relative reductions in sexually violent behavior were driven by changes in the key theoretical mediators such as knowledge of sexual violence legality and harm, and victim empathy (16).

This paper presents baseline data from faculty participating in the SCALE trial, which is designed to test implementation strategies for promoting engagement with, uptake of, and effectiveness of GlobalConsent. As noted, keys to intervention uptake are the KAB of University faculty and leaders at the participating Universities, as well as the beliefs about the acceptability and feasibility of the implementation. Prior to any implementation activities, faculty at the participating Universities completed an online survey that measured a range of KAB, and perceptions of the feasibility and acceptability of implementing sexual violence prevention programming at their University. This paper presents data from that faculty baseline survey with three goals:

1. To describe the KAB around sexual violence concepts, and perceptions of the acceptability and feasibility of implementing sexual violence prevention programming among faculty;
2. To examine variation in sexual violence KAB and implementation feasibility and acceptability by University, and faculty sex, age and rank;
3. To examine predictors of faculty perceptions of implementation acceptability and feasibility, including faculty demographics and KAB around sexual violence.

## Method

### Setting and Context

The data for the present analysis represent the baseline data from a parent implementation trial, called “SCALE: Strategies for Implementing GlobalConsent to Prevent Sexual Violence in University Men.” The purpose of the parent trial is to implement GlobalConsent at seven Universities of Medicine and Pharmacy across all regions of Vietnam, and to test two different packages of implementation strategies. As described elsewhere (23), the seven Universities are in the North (2), Central (2), and South (3) of Vietnam.

### Ethics review

The Institutional Review Boards at Emory University (STUDY00006481: SCALE) and the Hanoi University of Public Health (412/2023/YTCC-HD3) in Vietnam reviewed and approved the study.

### Sample and recruitment

The final sample for this paper consists of 2343 faculty from the seven participating Universities. All full-time permanent faculty (termed lecturers) who were currently working (i.e., not on extended leave) were invited to participate in the survey. Faculty were recruited for the study via email and text messages with a series of communications being sent by the local research partners. In addition, formal messages were sent to Faculty by the University and Departmental leadership prior to and during recruitment to endorse the study and to encourage faculty to respond to the survey. Reminders for survey completion were sent via REDCap, email, and text messaging to faculty at each institution. Response rates were consistent across universities with one exception. A total of 2343 of 3441 eligible faculty responded, for an overall participation rate of 68.1%. Response rates across Universities were as follows: University [U]1 = 72.5%; U2 = 78.8%; U3 = 99.1%; U4 = 79.4%; U5= 75.4%; U6 = 41.1%; U7 = 81.0%.

### Measures

All measures and scales were developed originally in English and then were translated to Vietnamese. All measures were subjected to cognitive testing to ensure they were sensible and captured the intended concept, and minor wording changes were made when needed. Translated scales are available in the published protocol (23). Unless otherwise indicated, all scale items were assessed on five-point Likert scales, where 1 = Totally Disagree, 2 = Disagree, 3 = Neutral, 4 = Agree, and 5 = Totally Agree.

*Demographic characteristics*. Several demographic characteristics were assessed including age, sex at birth, gender identity, ethnicity, current sexual orientation, number of years at the University, and current faculty rank. Questions on sex at birth, gender identity, and sexual orientation followed recommendations from the Nation Academies of Sciences, Engineering, and Medicine (47). Other demographic questions were based on those previously administered in the GlobalConsent efficacy trial (24).

*Acceptability and feasibility of sexual violence prevention programming* was assessed using Weiner and colleagues’ (48) measure. Weiner’s original scale included 12 items mapping onto three constructs of implementation (acceptability, appropriateness, and feasibility). After adjusting wording of the items to the Vietnamese language (23), ten items were included in the survey. A factor analysis showed two clear factors, one including the five items assessing acceptability and appropriateness (e.g., “Implementing sexual violence prevention programming with students at our university is something I support.”), and the other consisted of the five items assessing feasibility (e.g., “Implementing sexual violence prevention programming with students at our university would be easy to do”). Indices of Acceptability and Feasibility were computed by averaging the five items in each scale. In this sample, internal consistency of the acceptability/appropriateness scale was α = .91, and internal consistency of the feasibility scale was α = .90.

*Legal knowledge of sexual violence* was assessed using Legal Knowledge scale (49), which asks whether a series of behaviors are (1) Illegal, (2) Legal, but harmful, (3) Legal and not harmful. Items included four sexual acts done via force (e.g., “Forcing a person to have oral sex.”), and eight items describing sexual contact without force (e.g., “Unwanted hugging or kissing”, “Pressuring someone to have sex”). The Vietnamese version of this scale was used successfully in the GlobalConsent efficacy trial (16). Factor analyses confirmed that the two scales were distinct and correlated, so we averaged the responses for items loading on each factor to create two indices regarding perceptions of Illegality of sexual contact. Items were reverse scored prior to averaging so higher scores indicated a belief that the items are illegal. In this sample, internal consistency of the forced sexual violence was α = .79, and internal consistency of the non-forced, non-consensual sexual violence scale was α = .82.

*Active consent knowledge* (50) was assessed with 12 statements describing situations involving sexual consent were presented and respondents rated their agreement with those items. Items included general sexual situations (e.g.,” If a person consents for sex, one can continue sexual contact even if the person changes their mind”), situations focusing on casual vs. committed partners (e.g., “Obtaining consent for sex is just as necessary with a casual partner as it is with a committed partner”), and situations involving alcohol (e.g., “A woman who is drinking alcohol heavily can still give legal consent to sexual activity). The Vietnamese version of this scale was used successfully in the GlobalConsent efficacy trial (Yount et al. 2022). Factor analyses in the present sample did not yield conceptually distinct constructs; however, the 12 items had adequate internal consistency in the present sample (ɑ = .75), so a unidimensional scale was created by averaging the 12 items. Higher scores indicated a greater understanding of sexual consent.

*Rape myth acceptance* was assessed with 15 items taken from McMahon (51) and Lanier and Elliot (52) and adapted successfully for use in the GlobalConsent efficacy trial (Yount et al. 2022). Respondents rated their agreement or disagreement with each statement. Factor analyses yielded two distinct but correlated factors, one including 12 items focused on victim behavior (e.g., “If a girl doesn’t say “no” she can’t claim rape”, “If a woman dresses in a sexy dress, she is asking for sex”), and 3 items focuses on male or perpetrator behavior (e.g., “Rape happens when a guy’s sex drive gets out of control”). Two indices were created by averaging the 12 items focused on victim behavior (α = .90) and the 3 items focused on perpetrators (α = .79).

*Campus climate for sexual violence* was assessed with six items (53) that assessed agreement with the statement that students who reported being abused, sexually assaulted or stalked would be (1) believed, (2) respected, and (3) admired. The three items were asked with reference to “Most faculty at my University”, and to “Most leaders at my University”. Though items were asked separately in reference to University faculty and University leaders, a factor analysis indicated a single factor for all six items, as did the estimate for internal consistency (ɑ = .88) in this sample. Thus, the responses for all six items were averaged to create a single score of campus climate around sexual violence, with higher scored indicating greater support for student victims.

*Institutional norms about sexual violence*. Faculty responded to three statements that were developed for this study describing the problem of sexual violence (“Sexual violence is rare at my University”, “Sexual violence happens among students at my university but it is not a big problem”, “Sexual violence among students is an issue at other universities but not mine”). The items were not highly correlated, and internal consistency of the items was not adequate (ɑ = .43). Thus, the items were considered separately.

### Analytic plan

To address the aims of the study, we first computed descriptive statistics for the sample overall, and examined differences by key demographics including sex, age, and University. We then examined predictors of implementation acceptability and feasibility of sexual violence programming using linear regression modeling. First, bivariate linear regressions were estimated to assess the relationship of each implementation outcome with each demographic variable and each KAB variable separately. Then, multivariable linear models were estimated. We used a purposeful variable selection method (54, 55) to build multivariable models. Specifically we included key demographics in all models (i.e., University, Faculty Sex at birth, lecturer status, and Faculty age group) as covariates, and substantive KAB predictors that had significant bivariate relationships with the outcome at the *p* < .10 level. Time employed at the University was highly correlated with faculty age (*r* = .80), and so was not considered in the multivariable models. For all multivariable models, we examined collinearity diagnostics; in all models, all variance inflation factors were below 5 and all tolerance estimates were greater than 0.10, indicating no problematic collinearity.

## Results

### Descriptive data

The mean age of the sample was 38.0 with a range of 21-72 years. The sample was 59.2% female and 40.8% male according to sex at birth. Regarding gender identification, 59.2% identified as female, 40.8% as male, 0.1% transgender, and 0.1% as other. Almost 97% of the sample identified as “straight,” with the remaining 3% identifying as gay, bisexual or some other sexual orientation. Virtually all participants (97.9%) identified Kinh as their ethnicity. With regard to position rank, just over 79.3% of the sample reported their position as Assistant Lecturer or Lecturer, and 20.7% reported their position as Key or Senior Lecturer. Few participants (3.1%) reported any prior training in sexual violence prevention, and just over 7% had heard of the GlobalConsent program.

Means and standard deviations for all substantive variables are shown in Table 2 for the full sample and by University, including the p-value from an ANOVA comparing means by University. For perceptions of university sexual violence, the overall means were near the midpoint of the scale (3.68, 2.46, 2.96), indicated on the whole, faculty did not see sexual violence as a major problem at their university. Regarding faculty’s legal knowledge about sexual violence, means show that virtually all faculty endorsed forced sexual contact as illegal (M = 2.97 on a 1-3 scale), but non-forced non-consensual sexual contact items (M = 2.46) were rated between illegal, and ‘not-illegal but harmful.’ Regarding beliefs about active consent, the overall mean of 3.87 on a 5-point scale indicated that faculty endorsement of consent beliefs was moderate. Regarding Rape Myth acceptance, participants tended to disagree with victim-focused rape myths (*M* = 1.82 on a 5-point scale) but higher for perpetrator focused myths (*M* = 3.24 on a 5-point scale), where faculty were mostly neutral. Regarding perceptions of campus climate, faculty believed that both faculty and leaders at their university provided moderate strong levels of support for student victims of sexual violence (*Ms* = 3.96 and 3.92, respectively). Finally, faculty ratings regarding implementation of sexual violence prevention programming at their University for Acceptability (*M* = 4.2 on a 5-point scale) and Feasible (4.0 on a 5-point scale) were both high; comparing Acceptability and Feasibility means with a paired t-test showed that faculty saw implementation as more Acceptable than Feasible, *t* (2326) = 20.13, *p* < .001.

**Table 1.** Sample Characteristics: N and % or Mean and standard deviation, Baseline Faculty Survey of the SCALE Implementation Trial, 2024-2025.

|  | All Universities<br>N = 2343<br>68.1% | Uni 1<br>N = 605<br>72.5% | Uni 2<br>N = 328<br>78.8% | Uni 3<br>N = 216<br>99.1% | Uni 4<br>N = 377<br>79.4% | Uni 5<br>N = 309<br>75.4% | Uni 6<br>N = 383<br>41.1% | Uni 7<br>N = 124<br>81.0% | Test statistic |
| --- | --- | --- | --- | --- | --- | --- | --- | --- | --- |
| Sex at birth N (%) |  |  |  |  |  |  |  |  |  |
| Female | 1373 (59.2) | 56.2 | 68.5 | 79.5 | 56.5 | 47.5 | 56.7 | 58.5 | $\chi^2 (6) = 70.0, p < .001$ |
| Male | 945 (40.8) | 43.8 | 31.5 | 20.5 | 43.5 | 52.5 | 43.3 | 41.5 |  |
| Age, Mean (sd) | 38.0 (8.9)<br>range = 21-72 | 39.1<br>(8.2) | 36.6<br>(7.7) | 35.3<br>(8.4) | 36.7<br>(9.5) | 36.8<br>(8.5) | 39.6<br>(7.6) | 43.1<br>(13.7) | $F (6, 2259) = 18.2, p < .001$ |
| Ethnicity N (%) |  |  |  |  |  |  |  |  |  |
| Kinh | 2281 (98.0) | 98.2 | 99.4 | 99.5 | 99.7 | 96.4 | 95.8 | 95.9 | $\chi^2 (18) = 45.3, p < .001$ |
| Other | 47 (2.0) | 1.8 | 0.6 | 0.5 | 0.3 | 3.6 | 4.2 | 4.1 |  |
| Sexual orientation % |  |  |  |  |  |  |  |  |  |
| Straight | 2248 (96.8) | 99.0 | 98.5 | 96.3 | 97.9 | 93.1 | 94.4 | 95.9 | $\chi^2 (6) = 34.2, p < .001$ |
| Gay/Bisexual/other | 74 (3.2) | 1.0 | 1.5 | 3.7 | 2.1 | 6.9 | 5.6 | 4.1 |  |
| Years at Univ, Mean (sd) | 11.0 (7.9)<br>range = 0-42 | 10.9<br>(7.8) | 10.8<br>(6.7) | 10.2<br>(7.2) | 11.7<br>(8.7) | 11.1<br>(8.5) | 12.5<br>(8.1) | 5.9<br>(5.3) | $F (6, 2232) = 11.1, p < .001$ |
| Type of position N (%) |  |  |  |  |  |  |  |  |  |
| Assistant/Lecturer | 1796 (79.4) | 75.5 | 81.4 | 95.7 | 70.2 | 81.8 | 78.5 | 88.8 | $\chi^2 (6) = 66.5, p < .001$ |
| Key/Senior | 467 (20.6) | 24.5 | 18.6 | 4.3 | 29.7 | 18.2 | 21.5 | 11.2 |  |
| Had prior participation in sexual violence programming (%) | 3.1 (N=71) | 3.2 | 0.9 | 4.8 | 2.2 | 3.3 | 5.0 | 1.7 | $\chi^2 (6) = 13.8, p = .03$ |

**Table 2.**
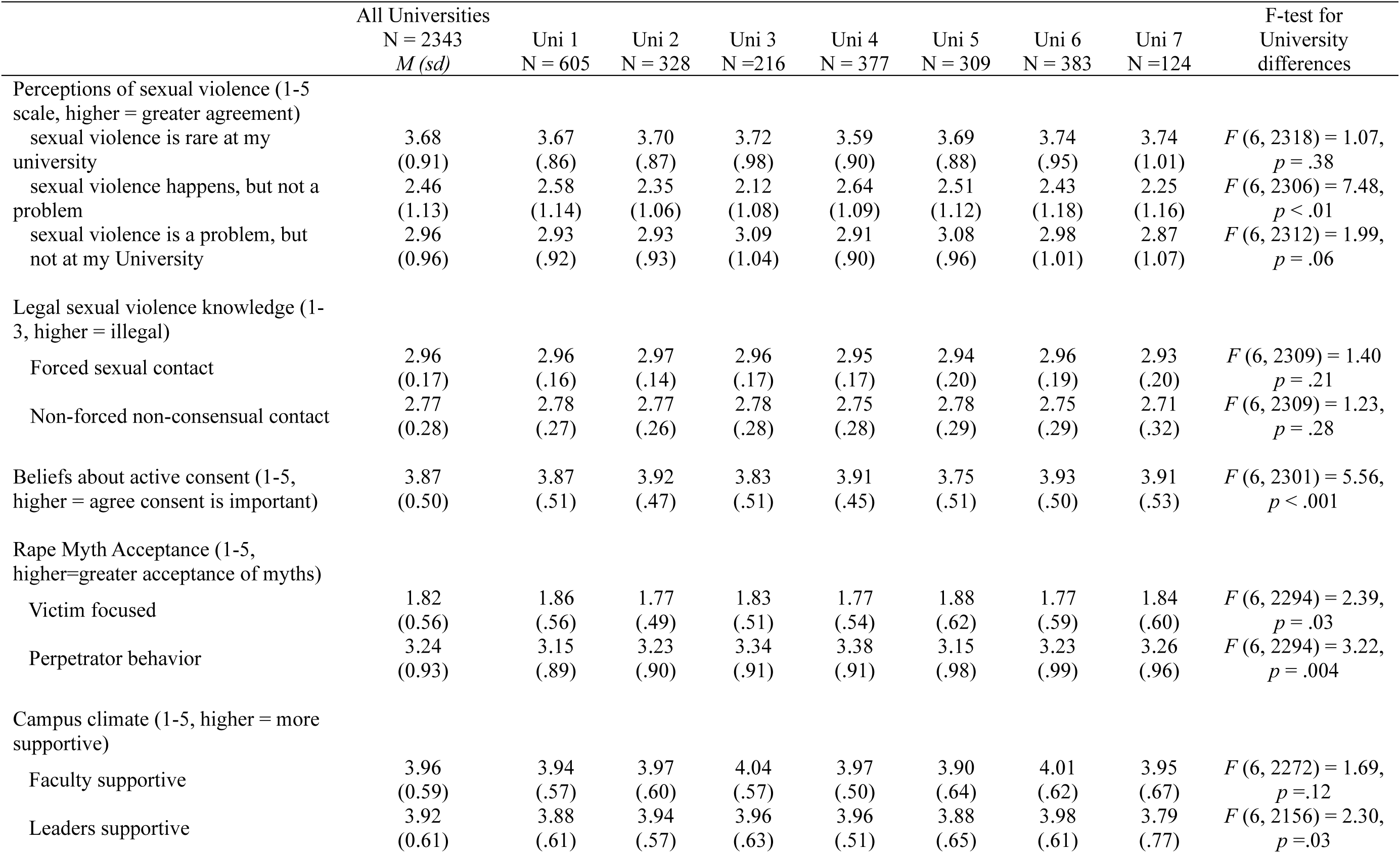
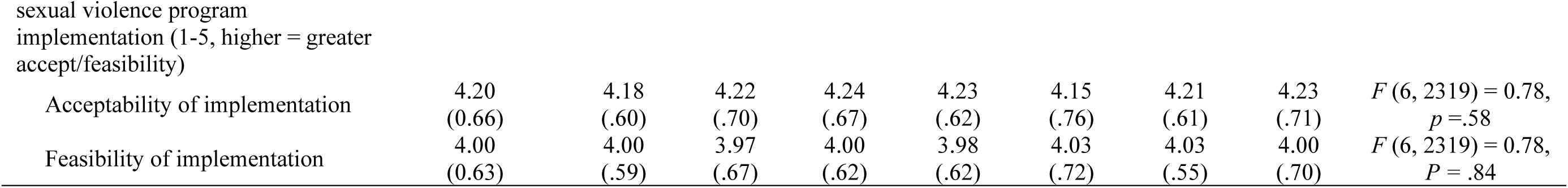
Means and standard deviations for KAB variables by university, Baseline faculty survey of the SCALE Implementation Trial, 2024-2025.

|  | All Universities<br>N = 2343<br>*M (sd)* | Uni 1<br>N = 605 | Uni 2<br>N = 328 | Uni 3<br>N = 216 | Uni 4<br>N = 377 | Uni 5<br>N = 309 | Uni 6<br>N = 383 | Uni 7<br>N = 124 | F-test for<br>University<br>differences |
| --- | --- | --- | --- | --- | --- | --- | --- | --- | --- |
| Perceptions of sexual violence (1-5<br>scale, higher = greater agreement) |  |  |  |  |  |  |  |  |  |
| sexual violence is rare at my<br>university | 3.68<br>(0.91) | 3.67<br>(.86) | 3.70<br>(.87) | 3.72<br>(.98) | 3.59<br>(.90) | 3.69<br>(.88) | 3.74<br>(.95) | 3.74<br>(1.01) | $F(6, 2318) = 1.07,$<br>$p = .38$ |
| sexual violence happens, but not a<br>problem | 2.46<br>(1.13) | 2.58<br>(1.14) | 2.35<br>(1.06) | 2.12<br>(1.08) | 2.64<br>(1.09) | 2.51<br>(1.12) | 2.43<br>(1.18) | 2.25<br>(1.16) | $F(6, 2306) = 7.48,$<br>$p < .01$ |
| sexual violence is a problem, but<br>not at my University | 2.96<br>(0.96) | 2.93<br>(.92) | 2.93<br>(.93) | 3.09<br>(1.04) | 2.91<br>(.90) | 3.08<br>(.96) | 2.98<br>(1.01) | 2.87<br>(1.07) | $F(6, 2312) = 1.99,$<br>$p = .06$ |
| Legal sexual violence knowledge (1-3, higher = illegal) |  |  |  |  |  |  |  |  |  |
| Forced sexual contact | 2.96<br>(0.17) | 2.96<br>(.16) | 2.97<br>(.14) | 2.96<br>(.17) | 2.95<br>(.17) | 2.94<br>(.20) | 2.96<br>(.19) | 2.93<br>(.20) | $F(6, 2309) = 1.40$<br>$p = .21$ |
| Non-forced non-consensual contact | 2.77<br>(0.28) | 2.78<br>(.27) | 2.77<br>(.26) | 2.78<br>(.28) | 2.75<br>(.28) | 2.78<br>(.29) | 2.75<br>(.29) | 2.71<br>(.32) | $F(6, 2309) = 1.23,$<br>$p = .28$ |
| Beliefs about active consent (1-5,<br>higher = agree consent is important) | 3.87<br>(0.50) | 3.87<br>(.51) | 3.92<br>(.47) | 3.83<br>(.51) | 3.91<br>(.45) | 3.75<br>(.51) | 3.93<br>(.50) | 3.91<br>(.53) | $F(6, 2301) = 5.56,$<br>$p < .001$ |
| Rape Myth Acceptance (1-5,<br>higher=greater acceptance of myths) |  |  |  |  |  |  |  |  |  |
| Victim focused | 1.82<br>(0.56) | 1.86<br>(.56) | 1.77<br>(.49) | 1.83<br>(.51) | 1.77<br>(.54) | 1.88<br>(.62) | 1.77<br>(.59) | 1.84<br>(.60) | $F(6, 2294) = 2.39,$<br>$p = .03$ |
| Perpetrator behavior | 3.24<br>(0.93) | 3.15<br>(.89) | 3.23<br>(.90) | 3.34<br>(.91) | 3.38<br>(.91) | 3.15<br>(.98) | 3.23<br>(.99) | 3.26<br>(.96) | $F(6, 2294) = 3.22,$<br>$p = .004$ |
| Campus climate (1-5, higher = more<br>supportive) |  |  |  |  |  |  |  |  |  |
| Faculty supportive | 3.96<br>(0.59) | 3.94<br>(.57) | 3.97<br>(.60) | 4.04<br>(.57) | 3.97<br>(.50) | 3.90<br>(.64) | 4.01<br>(.62) | 3.95<br>(.67) | $F(6, 2272) = 1.69,$<br>$p = .12$ |
| Leaders supportive | 3.92<br>(0.61) | 3.88<br>(.61) | 3.94<br>(.57) | 3.96<br>(.63) | 3.96<br>(.51) | 3.88<br>(.65) | 3.98<br>(.61) | 3.79<br>(.77) | $F(6, 2156) = 2.30,$<br>$p = .03$ |
| Acceptability of implementation | 4.20<br>(0.66) | 4.18<br>(.60) | 4.22<br>(.70) | 4.24<br>(.67) | 4.23<br>(.62) | 4.15<br>(.76) | 4.21<br>(.61) | 4.23<br>(.71) | $F(6, 2319) = 0.78,$<br>$p = .58$ |
| Feasibility of implementation | 4.00<br>(0.63) | 4.00<br>(.59) | 3.97<br>(.67) | 4.00<br>(.62) | 3.98<br>(.62) | 4.03<br>(.72) | 4.03<br>(.55) | 4.00<br>(.70) | $F(6, 2319) = 0.78,$<br>$P = .84$ |

We examined differences in KAB by University, Sex, Faculty Rank, and Age Group using one-way ANOVAs. Means and p-values from ANOVAs are displayed in Tables 2 and 3. Differences between Universities were found for some variables including for perceptions of sexual violence as a problem, beliefs about active consent, rape myths, and campus climate leader support. There were not clear patterns of differences across Universities however. For example, faculty at University 5 reported the lowest level of consent beliefs but also endorsed the fewest perpetrator focused rape myths. The differences across universities were mostly small in magnitude (.25 *sd* or less), but we included University in our multivariate regression models to control for any unmeasured variables at the institutional level.

**Table 3.** Means of KAB and implementation variables by sex, rank, and age group: Baseline faculty survey of the SCALE Implementation Trial, 2024-2025.

|  | Sex at birth |  |  | Faculty rank |  |  | Age group |  |  |  |  |
| --- | --- | --- | --- | --- | --- | --- | --- | --- | --- | --- | --- |
|  | Male | Female | F-test | Lecturer | Sr. Lecturer | F-Test | < 30 | 31-40 | 41-50 | 50+ | F-test |
| Perceptions of sexual violence |  |  |  |  |  |  |  |  |  |  |  |
| Sexual violence is rare at my University | 3.76<br>(.91) | 3.63<br>(.89) | $F(1, 2309) = 11.61, p < .001$ | 3.66<br>(.89) | 3.77<br>(.93) | $F(1, 2255) = 6.06, p = .01$ | 3.67<br>(.88) | 3.62<br>(.88) | 3.74<br>(.96) | 3.86<br>(.91) | $F(3, 2256) = 5.12, p = .002$ |
| Sexual violence not a problem at my University | 2.52<br>(1.16) | 2.41<br>(1.10) | $F(1, 2297) = 5.35, p = .02$ | 2.41<br>(1.10) | 2.65<br>(1.20) | $F(1, 2243) = 17.01, p < .001$ | 2.25<br>(1.07) | 2.43<br>(1.10) | 2.55<br>(1.18) | 2.80<br>(1.20) | $F(3, 2245) = 13.06, p < .001$ |
| Sexual violence problem elsewhere not here | 3.03<br>(.99) | 2.92<br>(.94) | $F(1, 2303) = 6.75, p = .009$ | 3.08<br>(.93) | 2.93<br>(.97) | $F(1, 2249) = 8.23, p = .004$ | 2.94<br>(.97) | 2.91<br>(.96) | 3.07<br>(.97) | 3.07<br>(.93) | $F(3, 2250) = 4.05, p = .007$ |
| Legal sexual violence knowledge |  |  |  |  |  |  |  |  |  |  |  |
| Forced sexual contact | 2.97<br>(.15) | 2.95<br>(.19) | $F(1, 2300) = 5.02, p = .03$ | 2.96<br>(.17) | 2.95<br>(.18) | $F(1, 2247) = 0.17, p = .67$ | 2.95<br>(.17) | 2.97<br>(.15) | 2.95<br>(.18) | 2.92<br>(.23) | $F(3, 2250) = 3.83, p = .009$ |
| Non-consensual non-forced contact | 2.78<br>(.27) | 2.76<br>(.29) | $F(1, 2300) = 2.54, p = .11$ | 2.76<br>(.28) | 2.78<br>(.28) | $F(1, 2247) = 4.02, p = .04$ | 2.72<br>(.29) | 2.77<br>(.27) | 2.80<br>(.27) | 2.73<br>(.32) | $F(3, 2250) = 8.21, p < .001$ |
| Beliefs about active consent | 3.82<br>(.51) | 3.91<br>(.49) | $F(1, 2291) = 18.704, p < .001$ | 3.88<br>(.50) | 3.86<br>(.51) | $F(1, 2238) = 0.41, p = .52$ | 3.87<br>(.47) | 3.93<br>(.51) | 3.82<br>(.49) | 3.76<br>(.49) | $F(3, 2241) = 10.24, p < .001$ |
| Rape Myth Acceptance |  |  |  |  |  |  |  |  |  |  |  |
| Victim focused | 1.96<br>(.59) | 1.72<br>(.52) | $F(1, 2284) = 101.59, p < .001$ | 1.80<br>(.55) | 1.88<br>(.58) | $F(1, 2232) = 9.09, p = .002$ | 1.72<br>(.55) | 1.78<br>(.56) | 1.87<br>(.57) | 2.02<br>(.52) | $F(3, 2236) = 17.41, p < .001$ |
| Perpetrator behavior | 3.33<br>(.90) | 3.18<br>(.95) | $F(1, 2284) = 15.89, p < .001$ | 3.25<br>(.92) | 3.20<br>(.97) | $F(1, 2232) = 1.12, p = .28$ | 3.19<br>(.96) | 3.26<br>(.92) | 3.15<br>(.87) | 3.43<br>(.93) | $F(3, 2236) = 4.87, p = .002$ |
| Campus climate |  |  |  |  |  |  |  |  |  |  |  |
| Faculty support | 3.95<br>(.58) | 3.97<br>(.60) | $F(1, 2262) = 0.73$<br>$p = .39$ | 3.97<br>(.59) | 3.92<br>(.56) | $F(1, 2210) = 2.87$<br>$p = .09$ | 4.00<br>(.59) | 3.99<br>(.58) | 3.92<br>(.60) | 3.86<br>(.58) | $F(3, 2213) = 4.84$<br>$p = .002$ |
| Leaders support | 3.95<br>(.61) | 3.91<br>(.62) | $F(1, 2147) = 1.96$<br>$p = .16$ | 3.92<br>(.62) | 3.90<br>(.57) | $F(1, 2098) = 0.32$<br>$p = .57$ | 3.95<br>(.64) | 3.93<br>(.60) | 3.89<br>(.60) | 3.88<br>(.61) | $F(3, 2100) = 1.03$<br>$p = .38$ |
| Implementation variables |  |  |  |  |  |  |  |  |  |  |  |
| Acceptability | 4.20<br>(.64) | 4.21<br>(.66) | $F(1, 2309) = 0.043$<br>$p = .85$ | 4.22<br>(.65) | 4.12<br>(.68) | $F(1, 2256) = 7.80$<br>$p = .005$ | 4.25<br>(.64) | 4.22<br>(.66) | 4.14<br>(.67) | 4.13<br>(.63) | $F(3, 2257) = 3.60$<br>$p = .01$ |
| Feasibility | 4.03<br>(.60) | 3.98<br>(.64) | $F(1, 2262) = 4.52$<br>$p = .03$ | 4.00<br>(.63) | 3.97<br>(.61) | $F(1, 2256) = 0.74$<br>$p = .39$ | 4.01<br>(.65) | 4.01<br>(.64) | 3.97<br>(.64) | 4.01<br>(.55) | $F(3, 2257) = 0.53$<br>$P = .66$ |

Differences by sex were found for most variables, with women reporting stronger agreement that sexual violence was a problem at their university, stronger agreement with statements about active consent, and endorsing fewer rape myths about victims and perpetrators. Interestingly, women reported implementation of sexual violence prevention efforts at their University as less feasible than men.

Differences in personal characteristics and KAB by faculty rank were found for many variables. Compared to senior lecturers, lecturers thought sexual violence was more of a problem at their University, held fewer victim-focused rape myths, and thought sexual violence programming was more acceptable. That said, lecturers also had poorer legal knowledge for non-consensual, non-contact sexually violent behaviors. There were no differences between the two faculty ranks for legal knowledge about forced sexual contact, beliefs about active consent, perpetrator focused rape myths, and feasibility of sexual violence programming.

Across age groups of participants, differences were also found for most variables. Though the patterns of means were not uniform across variables, younger faculty reported stronger belief that sexual violence was a problem at their University, fewer rape myths, a most positive campus climate from faculty for sexual violence victims, and believed sexual violence prevention efforts were more acceptable.

### Predicting implementation feasibility and acceptability

We next used linear regression models to examine the relationships of demographic and knowledge/attitude/belief variables with the two implementation outcomes of the feasibility and acceptability of sexual violence prevention. Results are shown in Table 4. For acceptability (column 2), bivariate regressions showed that younger age, rank as a more junior lecturer (vs. senior), and fewer years at the University were related to greater acceptability of sexual violence prevention programming. For KAB variables, several were related to the perceived feasibility of sexual violence prevention: endorsement that sexual violence is not a problem (negatively related), stronger agreement about the need for active consent, fewer victim focused rape myths, and stronger agreement that both faculty and leaders would support victims of sexual violence were all related to greater acceptability of sexual violence prevention programming. In the multivariable model predicting acceptability, the overall model was significant, *F* (16, 2040 = 6.44), *p* < .01, *R^2^*= .05. Several individual predictors were related to greater acceptability of sexual violence prevention: junior lecturer status (*b* = .09), stronger beliefs of the need for active consent (*b* = .08) and, belief that leaders support sexual violence victims (*b* = .18).

**Table 4.**
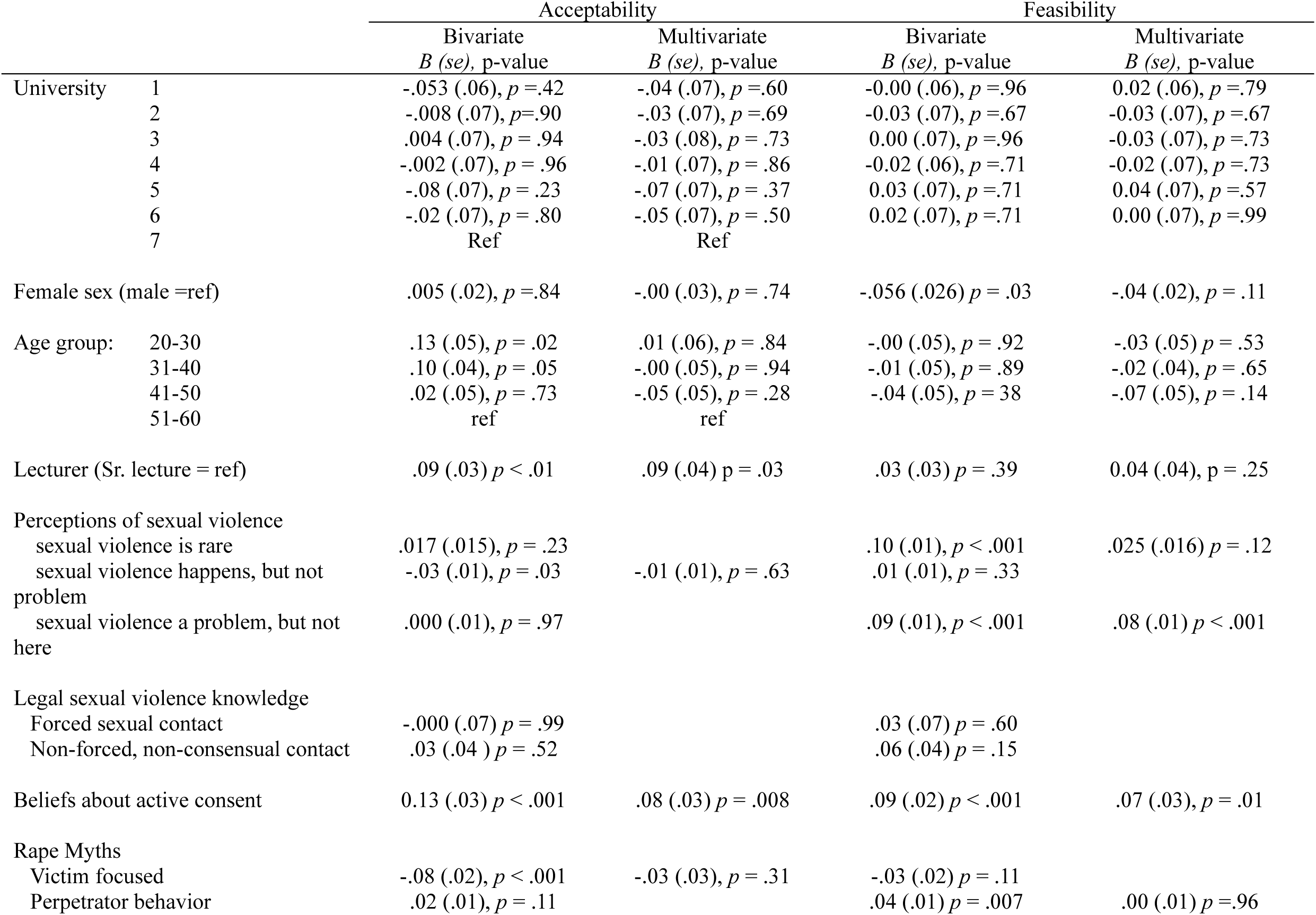
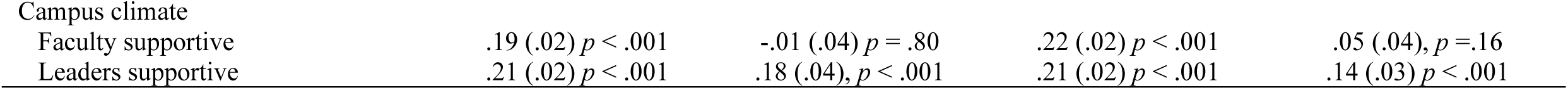
Bivariate and multivariate regression models predicting implementation acceptability and feasibility, Baseline faculty survey of the SCALE Implementation Trial, 2024-2025.

Results from bivariate and multivariate regression models are shown in the last two columns of Table 4. Bivariate analyses showed several variables related to stronger agreement about the feasibility of sexual-violence prevention programming: being female, beliefs that sexual violence is rare, beliefs that sexual violence is not a problem at the University, stronger beliefs about the need for active consent, greater endorsement of perpetrator-focused rape myths, and believing both faculty and leaders would support victims of sexual violence. The multivariable model predicting feasibility included nine variables and the overall model was significant, *F* (17, 2043 = 9.00), *p* < .001, *R^2^* = .06. Three individual-level predictors were related to greater perceived feasibility of sexual-violence prevention programming: beliefs that sexual violence is not a problem at the University (*b* = .08), stronger beliefs about active consent (*b* = .07), and stronger beliefs that University leaders would support victims of sexual violence (*b* = .14).

## Discussion

Sexual violence prevention programming has been strongly focused college campuses, and those efforts have primarily targeted students without engagement of University leaders and faculty, despite calls to do just that (19, 27, 56). Engaging faculty in sexual violence prevention on college campuses is important for several reasons. Faculty, as key constituents of the university setting, can contribute (favorably or unfavorably) to prevailing institutional norms about active consent and victim- and perpetrator-related myths about sexual violence. Faculty are often approached first by student victims of sexual violence, and the responses of faculty to these disclosures are key in promoting positive outcomes for those students. Finally, faculty knowledge, attitudes, and beliefs are instrumental in promoting the implementation of evidence-based prevention programming that is designed to engage students on campus.

This paper presents data from the first climate survey of faculty in Vietnam, completed as part of an implementation trial at seven Universities (23). To our knowledge, this is the first large scale survey designed to understand faculty KAB about sexual violence and prevention programming, with the goal of understanding how KAB among the faculty may affect GlobalConsent implementation and effectiveness (23). Specially, we examined faculty’s KAB about the prevalence and salience of sexual violence as a problem in university settings, their perceptions of the acceptability and feasibility of sexual violence prevention programming, and how KAB relates to the perceived acceptability and feasibility of sexual violence prevention programming on university campuses. Although GlobalConsent is designed to engage male students, understanding the KAB of influential organizational members and leaders where new evidence-based practices are being implemented has been shown to be an important driver of intervention uptake and sustainment (36, 38, 57). Universities are a key environment in which to implement sexual violence prevention programs for students (11, 19, 24, 27), so it is critical to understand faculty KAB around sexual violence, and their perception of sexual violence programs as they are implemented. This is particularly true in LMIC countries such as Vietnam, where cultural norms about sex and sexual violence are poorly documented, but cultural norms support sexual violence and sexual violence rates are high.

There are several key findings from this work. First, mean levels of faculty KAB regarding sexual violence concepts revealed some deficits that should be considered for future programming. On average, faculty failed to reject the statement that “sexual violence was rare at their University”, and did not strongly endorse the statement that “sexual violence is a problem at their university.” The reality, however, is that sexually violent behavior is fairly common; prior research in Viet Nam found a 6-month prevalence rate of sexually violent behavior of 30% among undergraduate men who participated in the GlobalConsent efficacy trial (15). Faculty also failed to strongly endorse beliefs about active consent, with a scale mean of 3.87 on a 5-point scale, a response of slightly less than ‘Agree’ for the ten statements regarding consent.

Rape myth focused on victims were largely rejected (mean of 1.8 on a 5-point scale), but myths regarding perpetrator behavior (e.g., that rape was driven by a high sex drive) were also not rejected, replicating prior work (45). On the positive side, statements about campus climate were largely positive, and faculty endorsed the notion (though not strongly) that sexual violence prevention programming was acceptable and feasible. Past research has identified potential barriers to implementation of sexual violence programming such as institutional culture, concern about reputational harm, and the belief that sexual violence was not common (also found here) (29).

There was some variation in these descriptive data, as would be expected, by demographic variables. Generally, female faculty saw sexual violence at the University as more common and problematic, held stronger beliefs about the importance of active consent, and reported fewer rape myths. They did not see campus climate for reporting sexual violence as more or less supportive than males, and there was no difference in the acceptability of sexual violence prevention programming. Ironically, female faculty reported sexual violence programming as less feasible than did male faculty, perhaps because they saw the problem as larger and more complex, or because of attitudes of fellow faculty. Likewise, faculty age was generally negative related to perceptions of the sexual violence are common and problematic, beliefs about active consent, and endorsement of rape myths. Younger faculty also reported higher levels of faculty support for victims (but not leader support), and thought sexual violence prevention programming was more acceptable.

Generally, faculty endorsed the acceptability and feasibility of sexual violence programming at their University with means slightly above a 4 on a 5-point scale. Bivariate predictors of acceptability and feasibility were examined, and several demographic variables and most KAB variables were related to acceptability and feasibility. However, in the multivariate context, two consistent predictors of acceptability and feasibility emerged: stronger beliefs about active consent was related to greater acceptability and feasibility, as was the perceptions that University leaders offer a supportive climate for sexual violence prevention. The latter variable – the extent to which leaders support a positive campus climate for sexual violence victims – was the strongest predictor of both acceptability and feasibility.

### Implications for implementation of prevention programming

One of the goals of the SCALE trial (23) is to test the implementation of several multilevel strategies that are intended to support the delivery and uptake of the GlobalConsent intervention, an efficacious web-based educational entertainment intervention that is tailored to heterosexual and bisexual male students in Vietnam (29). Those implementation strategies are designed to engage groups at different levels including the students, the local implementation support team, and the faculty and leaders at universities that are randomly assigned to more intensive community-engagement strategies. These data suggest that faculty-directed educational and engagement activities should address some of the aspects of faculty KAB revealed in preparatory key informant interviews (29) and in the survey findings presented here. Specifically, faculty perceptions of sexual violence as rare or not a problem at their university, endorsement of male perpetrator-focused rape myths, and weaker endorsement about active consent would be important targets for educational and norm-change efforts for faculty.

It is also important for university leaders to advance efforts that support a campus climate conducive to sexual violence reporting. The perceptions of leaders’ support for a positive campus climate around sexual violence prevention was the single strongest predictor of the perceived acceptability and feasibility of sexual violence prevention programming. In other words, for faculty to be on board with prevention efforts, it appears critical for leaders to demonstrate a positive campus climate that supports the victim-survivors of sexual violence. Given that faculty support and a positive climate for sexual violence prevention programming is key for implementation, training of university leaders may be a key in this process. Leaders can be trained and engaged to understand the importance of their role in foster positive faculty attitudes. Strategies such as the Leadership and Organizational Change for Implementation (LOCI) strategy are designed to train leaders to foster effective general leadership, implementation leadership, and a positive climate for implementation (58, 59). Implementation leadership is different from general leadership in that it focuses directly on perceptions of the importance of the new practice being implemented, and supporting the effective implementation of the new practice. Because faculty perceptions of both acceptability *and* feasibility of sexual violence prevention programming were related to leaders support, it appears important that leaders be trained both of fostering a positive climate around sexual violence programming (acceptability) and specific strategies to promote the prevention programming (feasibility)

### Study limitations

There are several limitations to the current study that should be acknowledged and provide opportunities for future research. First, the implementation outcome measures analyzed in the present studies were not translated and validated prior to our use in the current study. In particular, the measure of acceptability and feasibility was adapted from the measurement work of Weiner and colleagues (48) in which three concepts were developed: acceptability, appropriateness, and feasibility. Our adaptation of those items resulted in 12 items that yielded only two constructs in factor analysis.

Additionally, the focus of those items was on ‘sexual violence prevention programming”, a general topic rather than a specific intervention. As noted, this survey was part of a study testing implementation strategies for the GlobalConsent intervention, but too few faculty had heard of GlobalConsent at the time of the baseline survey to yield reliable data on perceptions of acceptability and feasibility of its implementation. Understanding the specifics of an intervention is critical for assessing its acceptability and feasibility as the fit between the intervention and context is seen as key for implementation success (60, 61). Opinions about the acceptability and feasibility of the GlobalConsent intervention may be quite different than general perceptions about sexual violence prevention. As the implementation trial continues, we will continue to collect data on the perceived feasibility and acceptability of GlobalConsent specifically from students, faculty, and leaders of participating universities.

A second limitation of the current analysis is that it employs cross sectional methodology, and all limitations of cross-sectional analyses apply. Most notably, we cannot make causal inferences about any of the relationship examined. For example, we do not know if the perceptions of campus climate as supportive drive implementation acceptability and feasibility, or the reverse. The longitudinal data collection planned in the parent study is designed specifically to overcome the limitation of unclear temporal ordering, and the randomized nature of the implementation trial will give us the future opportunity to assess the impacts of educational and norms-change outreach sessions with faculty to understand the impacts of these community-outreach efforts on the climate for sexual violence prevention among the faculty. Still, programming should seek to improve both campus climate and implementation perceptions.

Another potential limitation pertains to the representativeness of the sample. We achieved a 68% for a university-based survey, but our sample was slightly skewed, with female faculty more likely to participate than male faculty. Data obtained from the seven participating Universities indicated that, across all universities, 54.4% of all faculty are female. In our sample, 59.2% of baseline faculty survey participants were female. Given that sex differences exist in many of the substantive measures, with female faculty having more progressive attitudes, our results may not be an accurate reflection of population means at the participating universities.

This difference was most pronounced at university 6, where 47.4% of the faculty are female, but 56.7% of our sample was female (+9.3%). University 6 was also the largest University in the study and had the lowest response rate. It did not stand out as markedly different in other ways, however, including differences in faculty KAB. Despite this limitation, the participation rates observed in this study far surpass participation rates of faculty in climate surveys conducted in the United States (cites).

### Conclusions

This study is the first to conduct a large-scale assessment of Vietnamese faculty KAB around sexual violence and perceptions of implementation acceptability and feasibility of prevention efforts. Findings from the baseline survey of faculty across seven participating universities suggest that some KAB responses may support sexual violence (or may not strongly condemn it), but that faculty tended to perceive prevention programming as both acceptable and feasible. Multilevel intervention strategies targeting both students *and* University faculty and leaders may be needed to fully change campus climate and to support individual level interventions aimed at students. Faculty training may focus on changing KAB, and leader programming may focus on ensuring support for a positive campus climate – and making sure faculty are aware that leaders are supportive of sexual violence prevention. The SCALE trial (23) will implement leader and faculty-focused strategies in conjunction with a student-focused intervention, and will examine whether faculty KAB change, and how those changes relate to changes in student KAB. As many have argued (12, 19, 24, 27), sexual violence prevention efforts must include faculty and leaders to promote implementation of student directed programs and reduce sexual violence.

## Data Availability

The data will be made available upon conclusion of the trial in accordance with NIH guidelines for data access.

## Acknowledgement

This paper was supported by a research grant from the National Institute of Mental Health (NIMH; R01-MH133259; PI Yount), entitled *SCALE: Strategies for Implementing GlobalConsent to Prevent Sexual Violence in University Men*. The authors thank all implementation teams at participating universities in Vietnam for their assistance with data collection, and the implementing Universities for their support. The authors thank the following individuals for their assistance with portions of the project: Megan Sandal, Minh Anh, and Linh Kieu Pham. Finally, we thank all participants in the SCALE baseline faculty survey, without whom this analysis would not have been possible.

